# Assessment of Risk Factors Correlated with Age-Adjusted Mortality by Region for Prostate Cancer in Japan: an epidemiological study

**DOI:** 10.1101/2020.09.28.20164798

**Authors:** Takuya Nagano, Akihiko Hoshi

## Abstract

**Introduction:** Previous studies have suggested that smoking and drinking alcohol are prostate cancer risks. However, how regional unevenness of radiation oncologists/urologists in Japan or other risk factors including social background correlate with age-adjusted mortality of prostate cancer remain unknown.

**Methods:** In addition to known factors like smoking, we examined whether various other factors, such as the regional unevenness of radiotherapists/urologists, correlated with age-adjusted mortality rate (AAMR) for prostate cancer by prefecture. Statistical data were obtained from Annual Report of Hospital-Based Cancer Registries, Comprehensive Survey of Living Conditions and etc. We used the 10-year averages of AAMR from 2009 to 2018 in each prefecture to determine mortality rates.

**Results:** There was no correlation between the number of radiation oncologists/urologists per 100,000 and the 10-year averages of AAMR by prefecture in Japan. On the other hand, smoking rates and drinking per capita were correlated, respectively (P = 0.0002 and 0.0008).

**Conclusion:** The current study of statistics by prefecture in Japan also suggests that factors such as smoking and drinking alcohol are correlated with the risk of prostate cancer mortality. On the other hand, regional unevenness of radiation oncologists did not correlate. These results suggest that there is validity in this study design of analyzing factors of risk of prostate cancer mortality. And the study design was suggested to be useful for comparing various patient background factors.

## Introduction

With the rapid aging of the population, the number of prostate cancer patients in Japan is increasing rapidly[1-2]. The projected number of prostate cancer cases in 2019 was 78,500, making it the number one cancer case for men. And the projected number of deaths was 12,600 and the age-adjusted mortality rate (AAMR) was 2.2. [3]. Therefore, the question of how to detect and treat prostate cancer at an early stage has become an issue.

The incidence of prostate cancer has been reported to vary widely among ethnic, racial, and regional groups [4]. And the incidence of prostate cancer among Japanese immigrants to the United States was shown to be between that of Japanese and Americans [5]. Thus, it is suggested that not only genetic factors but also environmental factors increase the risk of morbidity. Environmental factors reported include lifestyle, metabolic syndrome, prostate inflammation, infection, prostate enlargement and chemical exposure [6]. Because there can be a variety of risk factors, it is difficult to determine that you had prostate cancer because of one factor. For example, with regard to lifestyle, smoking more often has been reported to increase the risk of prostate cancer in middle age [7-9]. The same is reported to be true for alcohol consumption [10]. In addition to these typical cancer risk factors, studies have examined whether prostate cancer is correlated with lifestyle factors such as high-fat diet, fatty fish consumption, milk consumption, fruit and vegetable consumption, physical activity, obesity, Japanese diet, Green tea and coffee consumption [11-26]. Prognostic factors may be considered in terms of lifestyle as well as social background. For instance, coffee consumption and poverty can be inversely correlated. And also poverty and suicide rates can be correlated [27, 28]. In more detail, coffee consumption can be correlated with social background factors such as suicide rates, amount of savings per family and unmarried rates. From the perspective of the biopsychosocial model, these social background factors need to be fully considered in epidemiology as well [29].

Health care is provided equitably to all prefectures in Japan. All Japanese people have access to health care without any disparity in wealth [30]. In addition, Japan is characterized by the fact that the country is subdivided into 47 prefectures. Each prefecture has its own characteristics, or trends, in the people of the county, and there are various statistics on these characteristics. This is called “Kenminsei” or characteristics of the people of a prefecture. This Keminsei causes differences in various prognostic factors in prostate cancer such as alcohol consumption, smoking rates, and even poverty in each region [31].

In the current study, we set up a study design to examine the contribution of risk factors and its associated social background factors including the regional unevenness of radiotherapists/urologists to the annual adjusted mortality rate of prostate cancer by region.

## Materials and Methods

Smoking and alcohol consumption have been suggested as mortality risks for prostate cancer in cohort studies [7-10]. In addition to these factors, we examined whether various other factors including social background correlated with AAMR for prostate cancer by prefecture. Statistical data were from the National Cancer Registry of Cancer Treatment Centers and Other Hospitals, the National Cancer Information Service, the National Cancer Institute Cancer Information Service, the Basic Living Survey, Vital Statistics, National Tax Administration statistics, the National Health and Nutrition Examination Survey, the Society of Radiotherapy, the Urological Society [32-38]. For mortality by prefecture, we used the mean of AAMR from 2009 to 2018. Factors for regional unevenness in the number of radiation oncologists per 100,000 and the number of urologists per 100,000 were calculated from the rosters of specialists published by the respective societies and the male population of the prefecture.

We included all prefectures in our analysis. However, only Kumamoto prefecture was excluded in the analysis of smoking rate because data on smoking rates were missing for this prefecture.

### Statistics

The correlations were tested by using the Pearson’s productive correlation coefficient. All the statistical analyses were performed using EZR software, version 1.37 (Saitama Medical Center, Jichi Medical University, Saitama, Japan), which is a graphical user interface for R (version 2.13.0; The R Foundation for Statistical Computing, Vienna, Austria). More precisely, it is a modified version of R commander (version 1.6-3), designed to include statistical functions that are frequently used in biostatistics. All the statistical analyses were two-sided, and a p value < 0.05 was considered statistically significant.

## Results

Smoking rates (p-value = 0.0002, Fig 1) and alcohol consumption per capita (liters) (p-value = 0.0008, Fig 2) were each correlated in 2017 for those aged 20 years and older.

**Fig 1.**
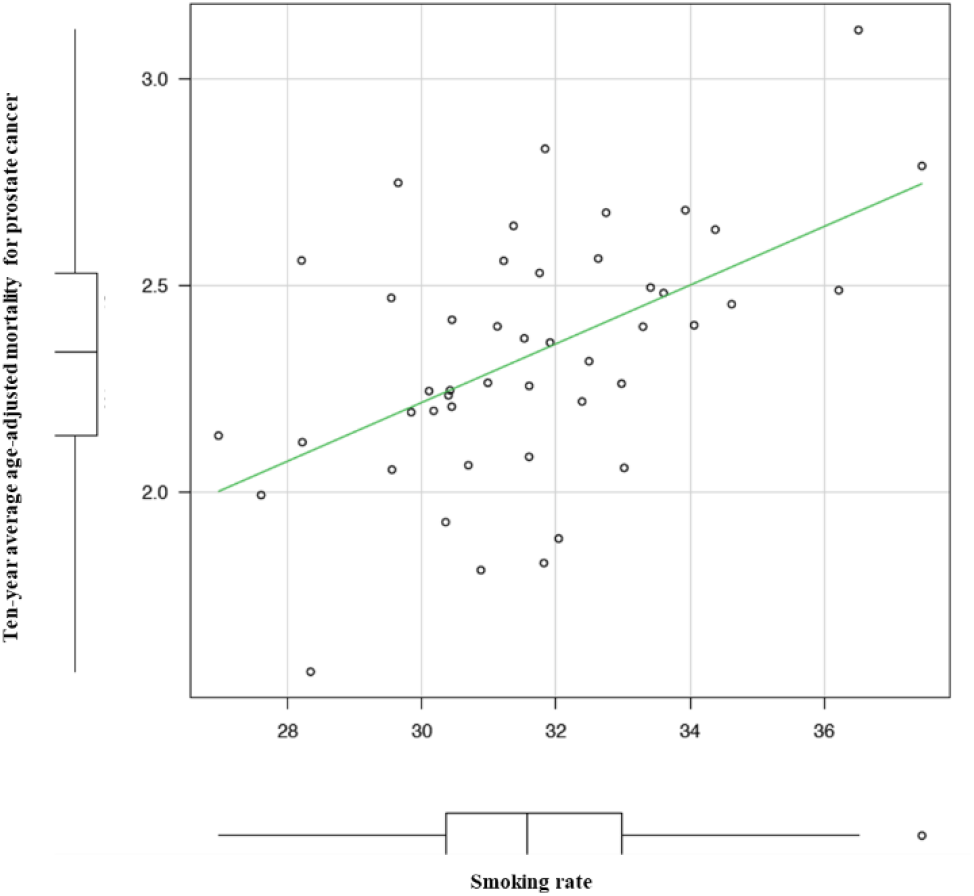
The correlation between smoking rate and Ten-year average age-adjusted mortality rate for prostate cancer by prefecture.

**Fig 2.**
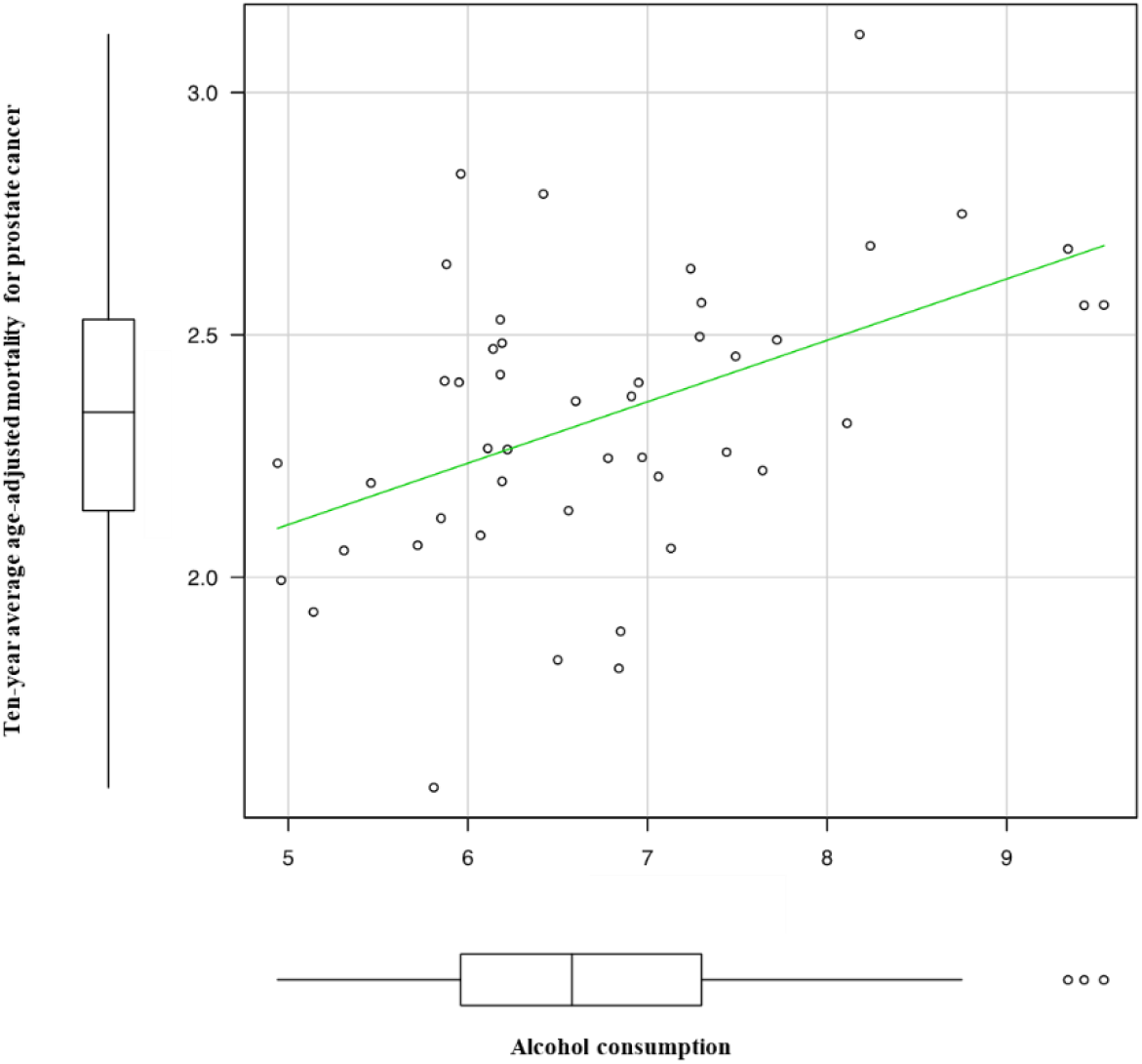
The correlation between alcohol consumption and Ten-year average age-adjusted mortality rate for prostate cancer by prefecture.

In addition, suicide rate (p-value = 0.015), household savings (p-value = 0.0007, negatively correlated), unmarried rate over 50 years of age (p-value = 0.0003), bread consumption (p-value = 0.001, negatively correlated), consumption of sunfish (p-value = 0.03), consumption of natto (p-value = 0.009), coffee consumption (p-value = 0. 03, negative correlation), and obesity rates (P-value = 0.006) were correlated (Table1).

**Table 1.** Relationship between ten-year average age-adjusted mortality rate for prostate cancer and various risk factors.

| Risk facotrs | Correlation coefficient | 95% CI | P-value |
| --- | --- | --- | --- |
| Smoking rate | 0.529 | 0.282-0.71 | 0.0002 |
| Alcohol consumption | 0.47 | 0.212-0.667 | 0.0009 |
| Suicide rate | 0.353 | 0.0732-0.581 | 0.02 |
| Household savings | -0.475 | -0.453 | 0.0007 |
| Unmarried rate over 50 years of age | 0.506 | 0.256-0.692 | 0.0003 |
| Bread consumption | -0.466 | -0.457 | 0.0010 |
| Mackerel pike consumption | 0.314 | 0.0298-0.552 | 0.03 |
| Natto consumption | 0.378 | 0.102-0.6 | 0.009 |
| Coffee consumption | -0.336 | -0.5145 | 0.02 |
| Obesity rate | 0.392 | 0.118-0.611 | 0.006 |

In contrast, there was no correlation between the number of radiation oncologists per 100,000 people, the number of urologists per 100,000 people, rate of PSA testing at health screenings in each prefecture in 2017, and the number of patients with stage IV prostate cancer according to cancer registry information. Other risk and preventive factors as referred to in the Japanese guideline did not correlate with seafood consumption, beef, chicken and pork consumption, vegetable consumption, milk consumption, green tea consumption, number of patients with hypertension, number of patients with diabetes, and sports population [6].

The correlation of the numbers between radiation oncologists and urologists per 100,000 people by prefecture is shown in Fig 3. There was a weak correlation between the number of radiation oncologists and the number of urologists, although not statistically different (P=0.06).

**Fig 3.**
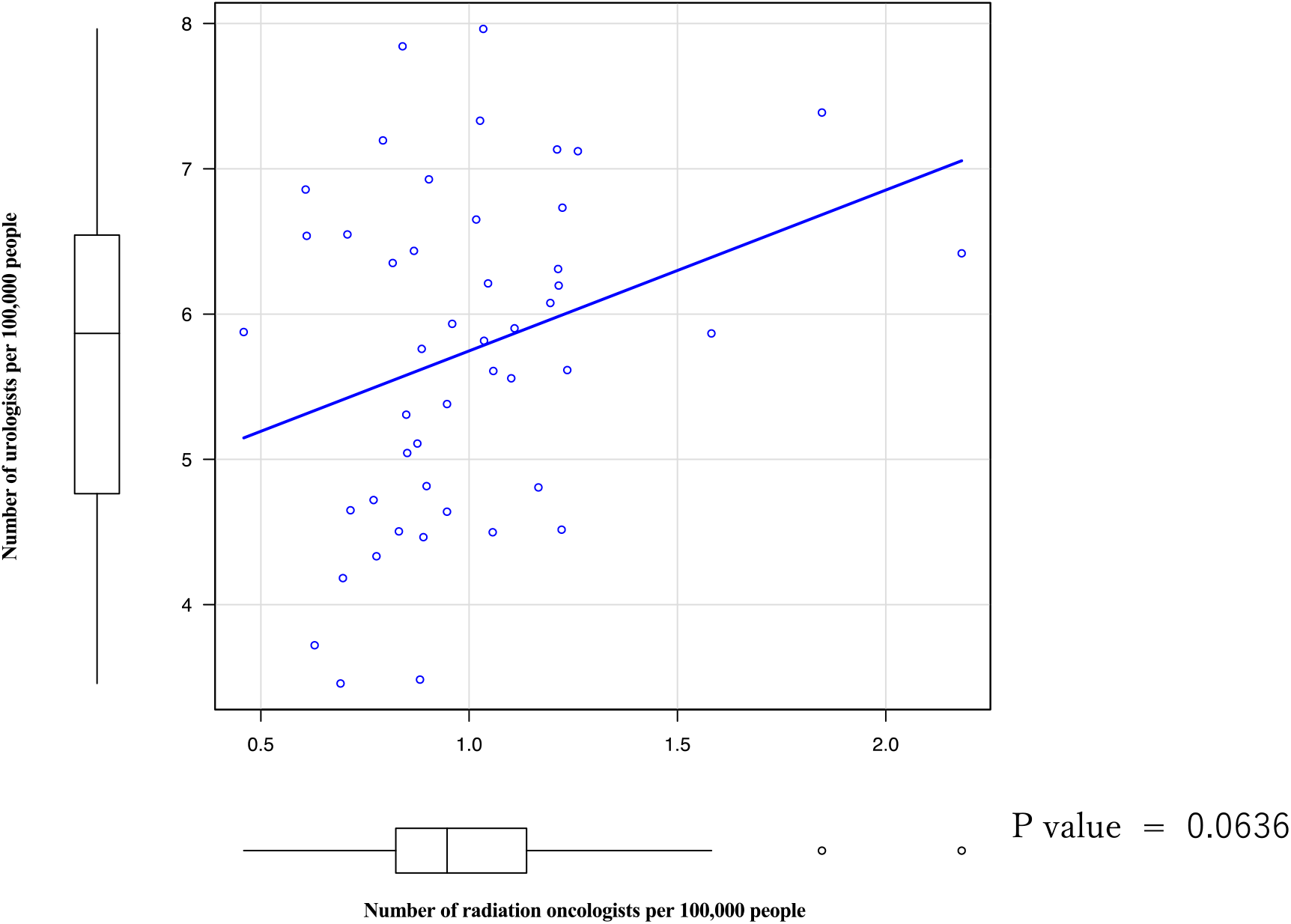
The correlation of the numbers between radiation oncologists and urologists per 100,000 people by prefecture

## Discussion

A study design to assess risk factors based on country- or region-specific morbidity and mortality and country-specific factors was recently conducted in the coronavirus disease 2019 (Covid-19) and BCG vaccination. However, this study has not been peer-reviewed, has confounding factors, and has not been scientifically evaluated [39]. On the other hand, this paper surprisingly led to randomized control trials and brought the mechanism of immunity in BCG into the limelight [40].

This study uses low-quality evidence observed at the population level to make broad inferences about the effectiveness of BCG at the individual level.

This epidemiological study has not been able to directly prove a relationship between BCG exposure rates and Covid-19 morbidity and mortality. This study was useful enough as a question to formulate a hypothesis. But this study should be treated as a hypothesis and not as evidence.

We used the same study design in our current study.

The statistics by Japanese prefectures also suggest that lifestyle and social background factors other than alcohol and smoking are associated with risk of prostate cancer mortality.

This study design was used for the first time in prostate cancer.

It may be useful in the epidemiology of prostate cancer to generate hypotheses, rather than as evidence, in the epidemiology of prostate cancer.

The current study suggests that, contrary to our expectations, the urban uneven distribution of physicians does not have a significant impact on patient outcomes. This suggests that the number of metastatic prostate cancer incidence does not correlate with the number of urologists and radiation oncologists.

The treatment of prostate cancer is well established for local treatment, and outcomes are excellent compared with other types of cancer. Rather, the problem is how to detect local prostate cancer at an early stage, rather than detecting it in its metastatic state.

Randomized control trials have already shown that as the screening exposure rate for PSA increases, the mortality rate decreases [41-43]. On the other hand, 10% of prostate cancers in clinical practice are found with bone metastases in Japan [44]. This suggests that screening rates in Japan are still at an inadequate level. There are also some factors that contribute to the disadvantage of PSA screening [45].

Although the goal of PSA screening for all men over 60 years of age could be targeted, the patient’s disadvantage needs to be taken into account. Therefore, the current study suggests the importance of intervening proactively with PSA screening according to the risks presented in the current study, i.e., depending on the patient’s background.

This has already been advocated for tailor-made screening; the EAU guidelines categorize risk according to age, family history, race, and previous PSA values [46].

As mentioned above, previous screening guidelines and previous papers have not included risk assessment of patients’ backgrounds.

However, clinicians could easily predict that low-income, obese, and unmarried men with drinking and smoking habits do not come to the clinic until they have symptoms of pain, which tends to delay detection. On the other hand, it is difficult to prove this prediction. This is because it is difficult to prove it without confounding factors. Therefore, we hypothesize that the current study will lead to the following: will there be a difference in mortality between prefectures that provide free, annual, and aggressive PSA screening for low-income obese unmarried men in their 50s who smoke or drink alcohol and prefectures that provide conventional, passive and optional PSA screening?

Whether tailor-made PSA screening by patient background risk factors can reduce mortality from prostate cancer should be studied prospectively.

## Data Availability

Availability of data and materials
The datasets used and/or analysed during the current study are available from the corresponding author on reasonable request.

## Acknowledgements

Not applicable.

## Availability of data and materials

The datasets used and/or analysed during the current study are available from the corresponding author on reasonable request.

## Consent for publication

Not applicable.

## Authors’ contributions

TN analyzed and interpreted the data. TN designed research. TN conducted data analysis. AH conducted review and editing. TN was a major contributor in writing the manuscript. All authors read and approved the final manuscript.

## Cosnflict of interest

Authors have nothing to declare.

The results of this research have not yet been presented at any conference.

